# Mapping of biomarker efficacy in SARS-CoV-2: tracking the impact of viral mutations and vaccinations

**DOI:** 10.1101/2022.12.03.22282974

**Authors:** ME Rahman Shuvo, Max Schwiening, Nikos Avramidis, Felipe Soares, Oliver Feng, Susana Abreu, Niki Veale, Q Gao, William Thomas, AA Roger Thompson, Richard J Samworth, Nicholas W Morrell, Kenneth Baillie, Stefan J Marciniak, Elaine Soon

**Author notes:** **Correspondence to:** Elaine Soon, Cambridge Institute for Medical Research, Keith Peters Building, Cambridge Biomedical Campus, Hills Rd, Cambridge CB2 0XY, UK. **Contributorship** MERS, NA, FS, WT, and NV collected data, reviewed source manuscripts, and helped analyze data. MS, FS, QG and SA contributed to the creation of online resources. OF and RJS performed statistical analyses. RT, NWM, SJM and KB contributed to the planning, organization, and funding of the study. ES conceptualized and organized the study, performed data analyses, funded the study, and takes responsibility for the integrity of the work as a whole. All authors contributed to the writing and critical revision of the manuscript. The corresponding author attests that all listed authors meet authorship criteria and that no others meeting the criteria have been omitted.

## Abstract

Rationale: Sophisticated prognostic scores have been proposed for SARS-CoV-2 but do not always perform consistently. We conducted these meta-analyses to uncover why and to investigate the impact of vaccination and variants.

Methods: We searched the PubMed database for the keywords “SARS-CoV-2” with “biomarker” and “mortality” for the baseline tranche (01/12/2020-30/06/2021) and either “SARS-CoV-2” or “Covid19” with “biomarker” and either “vaccination” or “variant” from 01/12/2020 to 31/10/2023. To aggregate the data, the meta library in R was used, and a random effects model fitted to obtain pooled AUCs and 95% confidence intervals for the European/North American, Asian, and overall datasets.

Results: Biomarker effectiveness varies significantly in different continents. Admission CRP levels were a good prognostic marker for mortality due to wild-type virus in Asian countries, with a pooled area under curve (AUC) of 0.83 (95%CI 0.80-0.85), but only an average predictor of mortality in Europe/North America, with a pooled AUC of 0.67 (95%CI 0.63-0.71, P<0.0001). We observed the same pattern for D-dimer and IL-6. This variability explains why the proposed prognostic scores did not perform evenly. Notably, urea and troponin had pooled AUCs ≥0.78 regardless of location, implying that end-organ damage at presentation is a key prognostic factor. The inflammatory biomarkers (CRP, D-dimer and IL-6) have generally declined in effectiveness in the vaccinated and variant cohorts. We note a significant lag from the pandemic advent to data availability and this has no doubt impacted on patient care.

Conclusions: Biomarker efficacies vary considerably by region. It is imperative that the infrastructure for collecting clinical data should be put in place ahead of a future pandemic.

## Disclosures

All authors have completed the ICMJE uniform disclosure form at http://www.icmje.org/disclosure-of-interest / and declare: no support from any commercial organisation for the submitted work; no financial relationships with any organisations that might have an interest in the submitted work in the previous three years; no other relationships or activities that could appear to have influenced the submitted work.

## Ethics approval

Ethical approval was obtained from the National Health System (UK) Health Research Authority via the Integrated Research Application System (reference 281880) for analysis of the Cambridge (UK) data. All the other data has been published and is in the public domain.

## Funding

ES and MS are supported by the UK Medical Research Council (MR/R008051/1); the British Medical Association (the Josephine Lansdell Award); and the Association of Physicians of Great Britain and Ireland (Young Investigator Award to ES); the Wellcome Trust ISSF and the Cambridge BHF Centre of Research Excellence (RE/18/1/34212). MES and WT are full-time NHS physicians who have volunteered their time for this work. FS received in-kind funding by the AWS Diagnostic Development Initiative and Google TPU Research Cloud. NV is supported by a BLF-Papworth Fellowship from the British Lung Foundation and the Victor Dahdaleh Foundation (VPDCF17-18). AART is supported by a British Heart Foundation Intermediate Clinical Fellowship (FS/18/13/33281). OF is funded by the StatScale programme (EP/N031938/1). RJS is supported by Engineering and Physical Sciences Research Council grants EP/P031447/1 and EP/N031938/1, as well as ERC Advanced Grant 101019498. SA and SJM are funded by the British Lung Foundation (VPDCF17-18), the Medical Research Council, UK (MR/V028669/1), the NIHR Cambridge Biomedical Campus (BRC-1215-20014) and the Royal Papworth NHS Trust. NWM is supported by the British Heart Foundation (SP/12/12/29836), the Cambridge BHF Centre of Research Excellence (RE/18/1/34212), the UK Medical Research Council (MR/K020919/1), the Dinosaur Trust, BHF Programme grants to NWM (RG/13/4/30107), and the NIHR Cambridge Biomedical Research Centre.

The funders did not have a role in the study design, data collection or analyses or the decision to submit for publication.

## Data sharing

The relevant anonymized data will be shared on reasonable request.

## To the Editor

SARS-CoV-2 is a novel beta coronavirus of zoonotic origin first identified in Wuhan, China at the end of 2019, which led to a Public Health Emergency of International Concern between February 2020 and May 2023. SARS-CoV-2 differs from previous viral threats in showing marked transmissibility during the asymptomatic/very early symptomatic stage(1) and person-to-person transmission by both airborne and fomite routes(2). At the beginning of the pandemic, there was no previous immunity, no known effective antiviral treatment, and no vaccine, resulting in a global death toll of >6.9 million (https://covid19.who.int/).

Due to the overwhelming number of cases and the significant morbidity and mortality associated with SARS-CoV-2, reliable prognostic scores are critically important to maximize survivorship. We noticed that some prognostic scores do not do well when applied away from their derivation region/population. For example, El-Solh(3) tested 4 peer-reviewed prognostic models constructed to predict in-hospital mortality for SARS-CoV-2 patients; proposed by Chen(4), Shang(5), Yu(6), and Wang(7). All the models examined had significantly worse validation area under curves (AUCs) than the area under curves of their derivation cohorts. For example, the AUC of the validation cohort using the model proposed by Chen(4) was at best 0.69 (95% confidence interval [CI] 0.66-0.72) compared to the derivation AUC, which was 0.91 (95% CI 0.85-0.97). We ran these meta-analyses to identify patterns in biomarker efficacy and to determine the effect of vaccination and virus variants on biomarker efficacy.

We searched the PubMed database for the keywords ‘SARS-CoV-2’ in combination with ‘biomarker name’ and ‘mortality’. The period for the first data tranche was set from 01^st^ December 2019 to 30^th^ June 2021. The second search covered 01^st^ December 2019 to 31^st^ October 2023 and used the following keywords:

a. ‘SARS-CoV-2’ with ‘vaccination’ and ‘biomarker name’
b. ‘SARS-CoV-2’ with ‘variant’ and ‘biomarker name’
c. ‘Covid19’ with ‘vaccination’ and ‘biomarker name’
d. ‘Covid19’ with ‘variant’ and ‘biomarker name’

All papers reporting mortality data for hospitalized patients swab-positive for SARS-CoV-2 with a biomarker level within 48h of admission were examined. Ethical approval was obtained from the Integrated Research Application System (reference 281880) for analysis of the Cambridge (UK) data. We excluded reports of patients already admitted to intensive care or restricted to specific groups (e.g. hemodialysis). Mortality (30-day or in-hospital) was used as the endpoint. This study is registered with PROSPERO (CRD42022366893); and the PRISMA flow diagram is shown in Fig.1(A). Further information is available at https://covid19.cimr.cam.ac.uk/.

**Figure 1.**
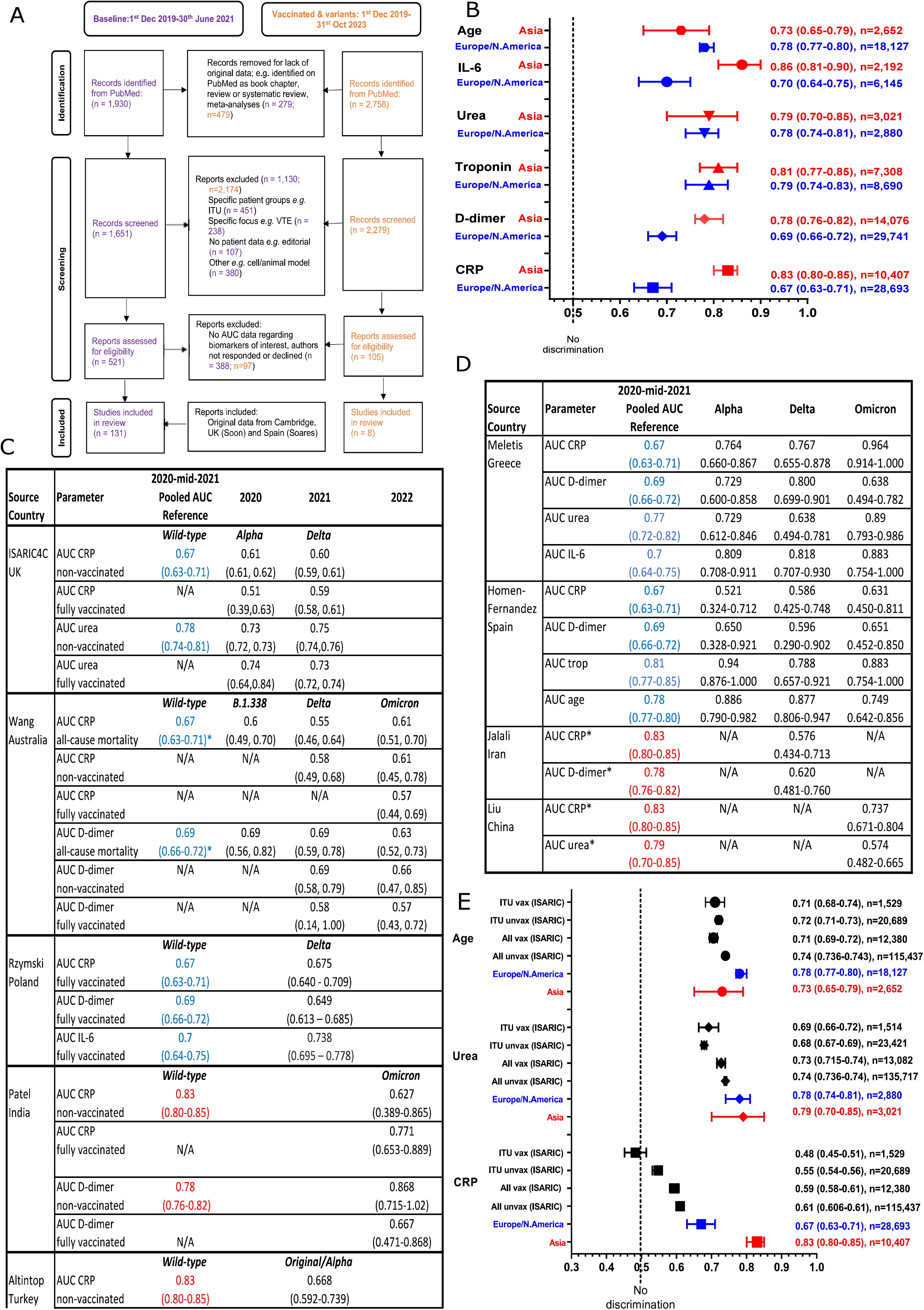
: Panel (A) shows a modified PRISMA flow diagram for paper review, selection, and inclusion in these meta-analyses for both the original/wild-type virus (in purple) and for variant and vaccination-related data (in orange). Panel (B) shows a summary forest plot from the meta-analyses of the first tranche of data (original/wild-type virus) with comparison of pooled area under curves for the five biomarkers being meta-analysed (CRP, D-dimer, troponin, urea, and IL-6) and age. Panel (C) shows results from variant cohorts, with the relevant pooled AUC from (A) as a reference. *indicate that the AUCs shown were for severe/critical illness rather than mortality. Panel (D) shows results from vaccinated cohorts, with the relevant pooled AUC from (B) as a reference. Panel (E) shows results from the ISARIC, GenOMICC and COG-UK consortiums; who have data relating to intensive care admissions and vaccination status. Throughout the panels, for ease of comparison, pooled values for Asian countries are shown in red and values for European/North American countries are shown in blue.

To aggregate the data on age and biomarkers from individual studies, the meta library in R was used to report overall mean values and 95% confidence intervals and the statistical significance of differences between mean values in the joint European and North American cohort and the Asian cohort. This analysis was based on estimates of standard errors for each study, obtained by assuming values for individual subjects were normally distributed in each study with a study-specific mean. In this way, measures of spread (IQR, SD and range) were converted into estimates of within-study standard deviations. Since the estimates of the study-specific means exhibited high levels of heterogeneity, a *random effects model* was fitted(8).

We examined 1,930 articles that were published from the beginning of the SARS-CoV-2 pandemic on 01^st^ December 2019 to 30^th^ June 2021 for the baseline study and 2,758 papers from 01^st^ December 2019 to 31^st^ October 2023 to obtain vaccination and variant data. The first phase meta-analyses revealed different patterns in the effectiveness of biomarkers in different regions of the world (Fig.1B). For example, admission CRP levels were a good prognostic marker for mortality in Asian countries, with a pooled AUC (area under curve) of 0.83 (95% CI 0.80-0.85) from 34 studies, but only an average predictor of mortality in Europe and North America, with a pooled AUC of 0.67 (95% CI 0.63-0.71, P<0.0001). This also held true for admission D-dimer and IL-6 levels. This could explain why the prognostic scores proposed for SARS-CoV-2 performed differently in different countries, as the ‘building blocks’ underpinning these prognostic scores have intrinsically different effectiveness in different populations. Interestingly, troponin and urea levels had universally ‘good’ pooled AUCs. This implies that end-organ damage at the time of presentation was a key prognostic indicator of severity for wild-type SARS-CoV-2 infection.

We expected that multiple rounds of vaccinations and ongoing mutations into different strains would significantly impact biomarker efficacy. The ‘inflammatory’ biomarkers (CRP, D-dimer and IL-6) have generally declined in effectiveness in the vaccinated and variant cohorts (Fig.1C-D). This is particularly illustrated by the Azarfar cohort(8) (Delta variant), who were unvaccinated, with AUCs of 0.576 (0.434-0.713) for CRP and 0.620 (0.481-0.760) for D-dimer. This is consistent with reports that the pathogenicity of SARS-CoV-2 has decreased in the order wild-type>Delta>Omicron and that the Omicron variant results in lower levels of IL-6 production(9, 10). It is also likely that the targeting of inflammation in general (e.g. dexamethasone) and IL-6 pathways in particular (e.g. tocilizumab) has had an effect on biomarker efficacy.

To examine this in greater detail we interrogated data collected from the ISARIC(11), GenOMICC(12) and COG-UK(13) consortiums. Fig.1(E) shows increasing loss of efficacy for CRP from the original meta-analyses to all unvaccinated in the cohort to all vaccinated to intensive care admissions. The AUCs for urea varied less between vaccinated and unvaccinated and had greater prognostic capability in every patient category examined, as did age. We think this shows that co-morbidity is now the leading factor in predicting susceptibility to fatal SARS-CoV-2 in the UK.

We hypothesize that biomarker efficacy in any population at this given time now hinges on many factors including:

A. Population factors – the numbers vaccinated and/or previously infected, the age and co-morbidities present,
B. Variant factors such as pathogenicity and transmissibility,
C. Socio-economic factors such as access to healthcare and nutritional status,

and that each country likely has its personalised ‘ingredient list’. From this it follows that prognostic biomarkers/scores should be individualized to particular populations. The simplest and most practical way to achieve this would be to set up information frameworks ahead of time, ideally online. To maximise efficiency, these processes should be automated as much as possible, e.g. by using programmes to harvest pre-specified data from medical software systems. Dunning *et al(14)* have proposed a multiple-tier system for this. We propose an even simpler set-up, with basic requirements being age, sex, biomarker levels, date of admission, date of outcome and outcome (death, survival, ITU admission and length of stay). Secondary outcomes (such as imaging data and complications) could be ‘bolt-on’ options. Furthermore, centres could be designated to collect data from specific groups, e.g. pregnant and immunocompromised people. In this way, data collection can be standardized rather than growing organically as was the case with the SARS-CoV-2 pandemic. The availability of such standardized data would facilitate rapid collation and meta-analyses. We note it took over a year for data collection for our initial tranche to be completed, by which time SARS-CoV-2 had evolved sufficiently for the original analyses to be outdated. This system would also allow the rapid organization of clinical trials by pinpointing ‘at-risk’ groups who could be targeted early for vaccination or intervention programmes. There are also significant geographical ‘black boxes’ where no published data is available. We would urge large organisations such as the World Health Organisation to identify this as an area requiring further funding.

## Data Availability

All data produced in the present study are available upon reasonable request to the authors.

## Acknowledgements

We thank Ms Natalie Doughty, Mr Chris Davies, Dr Benjamin Dunmore, and Mr Nikita Zubkov who provided general public and patient input into accessibility of data, website, and software.

Many thanks also to all root study authors, patients and essential workers who have selflessly shared their time and data.

## Notes

### Competing Interest Statement

The authors have declared no competing interest.

### Author Declarations

Ethical approval was applied for and approved by the National Health Service (UK) Health Research Authority and Health and Care Research Wales through the Integrated Research Application System (reference 281880) for analysis of the Cambridge (UK) data. The other data have been published and are in the public domain.

### Summary of Updates

Additional figure and text added to include vaccinated and variant data from the ISARIC, GenOMICC and COG-UK Consortiums.

## References (body of manuscript)

1. Yanes-Lane M, Winters N, Fregonese F, Bastos M, Perlman-Arrow S, Campbell JR, Menzies D. Proportion of asymptomatic infection among COVID-19 positive persons and their transmission potential: A systematic review and meta-analysis. PLoS One. 2020;15(11):e0241536.

2. Bak A, Mugglestone MA, Ratnaraja NV, Wilson JA, Rivett L, Stoneham SM, et al. SARS-CoV-2 routes of transmission and recommendations for preventing acquisition: joint British Infection Association (BIA), Healthcare Infection Society (HIS), Infection Prevention Society (IPS) and Royal College of Pathologists (RCPath) guidance. J Hosp Infect. 2021;114:79–103.

3. El-Solh AA, Lawson Y, Carter M, El-Solh DA, Mergenhagen KA. Comparison of in-hospital mortality risk prediction models from COVID-19. PLoS One. 2020;15(12):e0244629.

4. Chen R, Liang W, Jiang M, Guan W, Zhan C, Wang T, et al. Risk Factors of Fatal Outcome in Hospitalized Subjects With Coronavirus Disease 2019 From a Nationwide Analysis in China. Chest. 2020;158(1):97–105.

5. Shang Y, Liu T, Wei Y, Li J, Shao L, Liu M, et al. Scoring systems for predicting mortality for severe patients with COVID-19. EClinicalMedicine. 2020;24:100426.

6. Yu C, Lei Q, Li W, Wang X, Liu W, Fan X, Li W. Clinical Characteristics, Associated Factors, and Predicting COVID-19 Mortality Risk: A Retrospective Study in Wuhan, China. Am J Prev Med. 2020;59(2):168–75.

7. Wang K, Zuo P, Liu Y, Zhang M, Zhao X, Xie S, et al. Clinical and Laboratory Predictors of In-hospital Mortality in Patients With Coronavirus Disease-2019: A Cohort Study in Wuhan, China. Clin Infect Dis. 2020;71(16):2079–88.

8. Azarfar F, Abbasi B, Jalali A, Abbasian MH. Investigation of the relationship between monocyte chemoattractant protein 1 rs1024611 variant and severity of COVID-19. Cytokine. 2023;171:156367.

9. Barh D, Tiwari S, Rodrigues Gomes LG, Ramalho Pinto CH, Andrade BS, Ahmad S, et al. SARS-CoV-2 Variants Show a Gradual Declining Pathogenicity and Pro-Inflammatory Cytokine Stimulation, an Increasing Antigenic and Anti-Inflammatory Cytokine Induction, and Rising Structural Protein Instability: A Minimal Number Genome-Based Approach. Inflammation. 2023;46(1):297–312.

10. Korobova ZR, Arsentieva NA, Liubimova NE, Batsunov OK, Dedkov VG, Gladkikh AS, et al. Cytokine Profiling in Different SARS-CoV-2 Genetic Variants. Int J Mol Sci. 2022;23(22).

11. Garcia-Gallo E, Merson L, Kennon K, Kelly S, Citarella BW, Fryer DV, et al. ISARIC-COVID-19 dataset: A Prospective, Standardized, Global Dataset of Patients Hospitalized with COVID-19. Sci Data. 2022;9(1):454.

12. Pairo-Castineira E, Clohisey S, Klaric L, Bretherick AD, Rawlik K, Pasko D, et al. Genetic mechanisms of critical illness in COVID-19. Nature. 2021;591(7848):92–8.

13. Wright DW, Harvey WT, Hughes J, Cox M, Peacock TP, Colquhoun R, et al. Tracking SARS-CoV-2 mutations and variants through the COG-UK-Mutation Explorer. Virus Evol. 2022;8(1):veac023.

14. Dunning JW, Merson L, Rohde GGU, Gao Z, Semple MG, Tran D, et al. Open source clinical science for emerging infections. Lancet Infect Dis. 2014;14(1):8–9.

